# Developing and validating a machine learning model to predict successful next-day extubation in the ICU

**DOI:** 10.1101/2024.06.28.24309547

**Authors:** Samuel W Fenske, Alec Peltekian, Mengjia Kang, Nikolay S Markov, Mengou Zhu, Kevin Grudzinski, Melissa J Bak, Anna Pawlowski, Vishu Gupta, Yuwei Mao, Stanislav Bratchikov, Thomas Stoeger, Luke V Rasmussen, Alok N Choudhary, Alexander V Misharin, Benjamin D Singer, GR Scott Budinger, Richard G Wunderink, Ankit Agrawal, Catherine A Gao, the NU SCRIPT Study Investigators

## Abstract

**Background:** Criteria to identify patients who are ready to be liberated from mechanical ventilation are imprecise, often resulting in prolonged mechanical ventilation or reintubation, both of which are associated with adverse outcomes. Daily protocol-driven assessment of the need for mechanical ventilation leads to earlier extubation but requires dedicated personnel. We sought to determine whether machine learning applied to the electronic health record could predict successful extubation.

**Methods:** We examined 37 clinical features from patients from a single-center prospective cohort study of patients in our quaternary care medical ICU who required mechanical ventilation and underwent a bronchoalveolar lavage for known or suspected pneumonia. We also tested our models on an external test set from a community hospital ICU in our health care system. We curated electronic health record data aggregated from midnight to 8AM and labeled extubation status. We deployed three data encoding/imputation strategies and built XGBoost, LightGBM, logistic regression, LSTM, and RNN models to predict successful next-day extubation. We evaluated each model’s performance using Area Under the Receiver Operating Characteristic (AUROC), Area Under the Precision Recall Curve (AUPRC), Sensitivity (Recall), Specificity, PPV (Precision), Accuracy, and F1-Score.

**Results:** Our internal cohort included 696 patients and 9,828 ICU days, and our external cohort had 333 patients and 2,835 ICU days. The best model (LSTM) predicted successful extubation on a given ICU day with an AUROC 0.87 (95% CI 0.834-0.902) and the internal test set and 0.87 (95% CI 0.848-0.885) on the external test set. A Logistic Regression model performed similarly (AUROC 0.86 internal test, 0.83 external test). Across multiple model types, measures previously demonstrated to be important in determining readiness for extubation were found to be most informative, including plateau pressure and Richmond Agitation Sedation Scale (RASS) score. Our model often predicted patients to be stable for extubation in the days preceding their actual extubation, with 63.8% of predicted extubations occurring within three days of true extubation. We also tested the best model on cases of failed extubations (requiring reintubation within two days) not seen by the model during training. Our best model would have identified 35.4% (17/48) of these cases in the internal test set and 48.1% (13/27) cases in the external test set as unlikely to be successfully extubated.

**Conclusions:** Machine learning models can accurately predict the likelihood of extubation on a given ICU day from data available in the electronic health record. Predictions from these models are driven by clinical features that have been associated with successful extubation in clinical trials.

## Introduction

Mechanical ventilation is a life-saving intervention to support patients with respiratory failure. However, mechanical ventilation is also an invasive therapy with substantial risks.^1^ Hence, ICU physicians seek to extubate patients at the earliest point in their hospital course when they can sustain spontaneous breathing without an artificial airway.

Premature extubation can lead to reintubation, which is associated with a prolonged ICU stay and greater hospital mortality.^2^ As a result, physicians often delay weaning, prolonging the duration of mechanical ventilation and its associated complications.^3^ Implementation of protocolized daily screening by nurses or respiratory therapists followed by spontaneous breathing trials (SBTs) in the absence of physician input reduces the duration of mechanical ventilation and ICU stay.^4^ Despite this, compliance with these protocols is highly variable and SBTs alone have poor operating characteristics in predicting successful extubation.^5–8^ Furthermore, these protocols distract from other processes of care, increase costs, and are difficult to continue when ICU resources become limited.^9^

The success of protocol-driven weaning highlights the value of objective and data-driven approaches to assist healthcare providers identify candidates for liberation from mechanical ventilation. Electronic Health Records (EHRs) ingest continuous clinical data that report on patient physiology, including vital signs, laboratory results, medications, and ventilator parameters.^10^ Machine learning uses computational techniques that identify complex non-linear relationships within disparate data, including those in the EHR to improve the accuracy of clinical decisions.^11,12^ We sought to determine whether machine learning approaches applied to EHR data could predict successful next-day extubation. We trained our model using data from a relatively small cohort of well-phenotyped patients enrolled in an observational clinical cohort study. After careful annotation and labeling by clinicians, several models were found to perform well, accurately predicting successful extubation. Predictions from our models were driven by clinical features validated in clinical trials of protocol driven weaning. Our best performing model correctly predicted next-day extubation status in 84.6% of cases and could potentially have identified 35.4% (17/48) of failed extubation cases as unlikely to be successfully extubated.

## Methods

### Study participants and setting

Patients were enrolled in the Successful Clinical Response In Pneumonia Therapy (SCRIPT) Systems Biology Center, a single-site prospective cohort study of patients requiring mechanical ventilation, who underwent bronchoalveolar lavage for known or suspected pneumonia in a quaternary care hospital from 2018 to 2023 (NU IRB # STU00204868). Patients or their legal authorized representative consented to participate in this study. External testing was done using EHR data collected from patients in a mixed medical surgical ICU from a community hospital, Central DuPage Hospital (CDH) from 2018 to 2022 (NU IRB # STU00216678); this was a retrospective data-only protocol and received a waiver of informed consent. We included patients who required mechanical ventilation, with ICD-9 or ICD-10 codes for pneumonia or from the top 10 ICD codes of SCRIPT patient admissions (Supplemental Table 1); we excluded patients admitted from an operating room to minimize surgical ICU patients.

### Data

EHR data from both hospitals were extracted from the Northwestern University Enterprise Data Warehouse,^13^ and were manually reviewed and validated by ICU physicians who focused specifically on ventilation features and markers of intubation and extubation. Over two hundred charts were manually reviewed by physicians to ensure the accuracy of the extracted data. When the reviewing clinician was uncertain, a study adjudication committee meeting comprised of five critical care physicians made a consensus decision that was considered final. Our group is experienced in the annotation of clinical data and has adjudicated over 800 pneumonia episodes in a rigorous, predefined process.^14^ Each patient chart was further systematically reviewed by the research study team to gather information including transition to comfort measures only (CMO). For patients who had multiple sequences of intubation and extubation, we used only the first intubation/extubation sequence. We excluded patients with tracheostomy and failed extubation (requiring reintubation within two days) and excluded ICU days with palliative extubation and extubation while on extracorporeal membrane oxygenation (ECMO) from our training data.

### Features

Our features were inspired by the practice of daily multidisciplinary rounds in the ICU and built off a previously described dataset from our group.^15,16^ As almost all extubations occurred after 8AM (Supplemental Figure 1), we used only data from midnight to 8AM. This strategy is consistent with the protocol-driven weaning shown to reduce the duration of mechanical ventilation^4^ and can facilitate future deployment by presenting model predictions to the clinical team during morning rounds. For a given timestep, the model was fed with a dataset of 37 features pertaining to a specific patient, aggregated from the hours of 12AM-8AM from a single day. Features included in the dataset span ventilator parameters, laboratory values, mental status assessments, and organ failure assessments (Supplemental Table 2).

### Missing data

Missing data is common in real-world EHR data. Some missing data in the EHR are not random, instead reflecting a change in clinical status that results in different patterns of monitoring and documentation. We report the percent missing for each feature in Supplemental Table 2. Some machine learning models (such as XGBoost and LightGBM, detailed below and in Supplemental Methods) are designed to incorporate missing data. For other models, including RNN and LSTM, we tested several imputation strategies, including mean imputation and a binning strategy inspired by FIDDLE.^17^ For binning, features are coded into a flag indicating missing/present, and flags indicating the quartile range into which the data point falls. For example, a day with a respiratory rate of 24 breaths per minute would have a 0 for the missing flag (indicating the data are present), a 0 for the first bin that encompasses range <18 breaths per minute, a 0 for the second bin that encompasses range [18,22), a 1 for the third bin that encompasses range [22,27.4), and a 0 for the fourth bin that encompasses the highest range of ≥ 27.4. Details of each strategy are outlined in Supplemental Methods and in our code repository.

### Labels

We labeled the patient’s intubation status on each day as intubated, extubated, failed extubation, comfort measures palliative extuation, or pre-intubation. After removing pre-intubation days, and sequences where there was failed extuation or palliative extubation, our dataset contained only intubated days for training. The label of interest that our models predicted was the next day’s intubation/extubation status. Additional details on labeling and filtering are available in Supplemental Methods.

### Split

Our training dataset contains patients with hospital admission dates ranging from June 2018 to August 2023 (Figure 1). We split patients into train and test sets based on a cutoff date of August 20, 2021. This split put 80% of patient ecounters before that date into the train set, and the 20% after into the test set. We further split the train data into smaller train and validation subsets based on an 80/20% random split.

**Figure 1.**
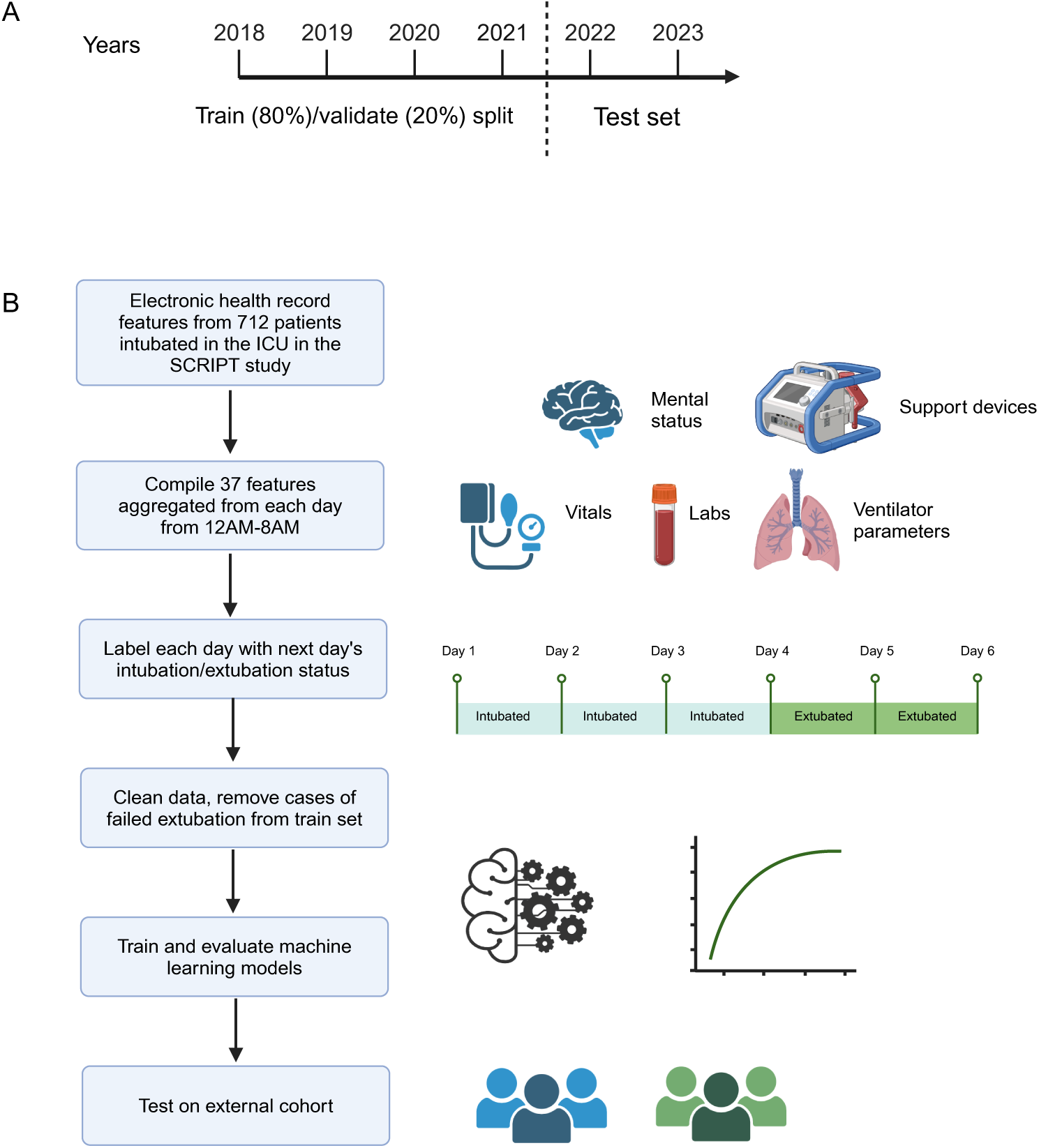
**(A) Data Split for Model Development:** Schematic describing the data split used in our clinical machine learning study. The dataset was divided into three distinct subsets: the training set, validation set, and test set. The training set was used to train the machine learning models, the validation set for tuning and model selection, and the test set to assess the final model’s performance and generalization to unseen data. **(B) Data processing pipeline;** full details are available in Supplemental Methods. Individual days are labeled as intubated or extubated, with 12AM-8AM aggregated features used to predict next-day intubation/extubation status. Data are cleaned, detailed in Methods and Supplemental Methods, before machine learning models are trained and evaluated.

### Modeling

Both traditional machine learning models and deep learning architectures were employed. We used traditional models, including logistic regression,^18^ random forest,^19^ LightGBM,^20^ and XGBoost,^21^ to make daily predictions. In contrast, deep learning models, specifically Recurrent Neural Networks (RNNs)^22^ and Long Short-Term Memory networks (LSTMs),^23^ used the entire patient sequence to make predictions. Detailed explanations and comparisons of the models are available in Supplemental Methods.

### Performance metrics

The primary metric for model evaluation was the Area Under the Receiver Operating Characteristic curve (AUROC). Additional metrics included Area Under the Precision-Recall Curve (AUPRC), Accuracy, F1-Score, Precision (PPV), Recall (Sensitivity), and Specificity.

### Feature importance

To understand and interpret the effect of the various features on the predictions made by our deep learning models, we used ablation techniques by masking individual features for 37 iterations (total features) and observing the decrease in AUC without that feature available. We also used XGBoost’s SHAP (Shapley Additive exPlanations) values.^24^ For our logistic regression model, we plotted the top feature coefficients. More detailed explanations are available in Supplemental Methods.

### Predicting failed extubations and missed opportunities for extubation

We defined a correct prediction of failed extubation as a patient who required reintubation within two days and a model prediction of a low probability of successful extubation on the day of extubation. We defined a possible missed opportunity for extubation as a patient with a high probability of successful extubation in the days before successful extubation.

### Statistics

Descriptive statistics are reported for the train/validate/test cohorts using % and median [Q1,Q3]. Comparisons between nonparametric data were done using Mann-Whitney U tests.

### Code and data availability

A detailed description of data extraction, processing, and modeling are available in our code repository at https://github.com/NUPulmonary/2024_Fenske_Peltekian. Programming was done in Python version 3.10. A deidentified version of the SCRIPT cohort data are available on PhysioNet, and the exact data needed to replicate this study will be posted in a future update.^16^

### Reporting checklist

We followed the TRIPOD Checklist for predictive model development, which is available in Supplemental Materials.

## Results

### Cohort description

There were 712 enrollments in SCRIPT during the study period, with 940 separate ICU stays totaling 16,402 ICU days. After filtering days and stays for our next-day successful extubation task, we trained and evaluated using 696 unique patients, 781 ICU stays, and 9,828 ICU days. Our CDH dataset consisted of 459 unique patients, 518 ICU stays, and 5,814 ICU days. This was filtered down to 333 unique patients, 349 ICU stays, and 2,835 ICU days. The failed extubation rate, defined as requiring reintubation within two days was 23.1% and 15.8% in the SCRIPT and CDH cohorts, respectively. Patient demographics and outcomes are reported in Table 1 between the train/validate/test and external sets. In the SCRIPT cohort, the median [Q1,Q3] patient age was 63 [51, 72], and 44% of the patients were female. The percent of patients with unfavorable outcomes (death, discharge to hospice, or lung transplantation) for the SCRIPT cohort was 45%. While the demographics and overall outcomes were similar among the different datasets, the features were markedly different (Table 2, Supplemental Table 2), likely reflecting changes in the cohort of patients requiring mechanical ventilation during the COVID-19 pandemic. A summary of the different types of intubation/extubation sequences and how often they occurred are presented in Supplemental Figure 2.

**Table 1.** Demographics and outcomes: Patient demographics and clinical outcomes of patients used for model training and testing, stratified across the training, validation, test, and external datasets. Continuous variables are presented as median [Q1,Q3]. Categorical variables are presented as number (percentages). Unfavorable outcome indicates a discharge to hospice or death. Except for four values of missing BMI, there were no other missing data.

|  | Train | Validate | Test | External |
| --- | --- | --- | --- | --- |
| Number of patients | 288 | 71 | 81 | 337 |
| Age, median [Q1,Q3] | 65.0 [52.0,73.0] | 59.0 [48.0,67.0] | 62.0 [49.0,71.0] | 64.0 [55.0,73.0] |
| Female, n (%) | 127 (44.1) | 31 (43.7) | 36 (44.4) | 130 (38.6) |
| Race, n (%) |  |  |  |  |
| Asian | 7 (2.4) | 1 (1.4) | 2 (2.5) | 16 (4.7) |
| Black or African American | 57 (19.8) | 14 (19.7) | 15 (18.5) | 22 (6.5) |
| Unknown or Not Reported | 50 (17.4) | 8 (11.3) | 15 (18.5) | 21 (6.2) |
| White | 174 (60.4) | 48 (67.6) | 49 (60.5) | 278 (82.5) |
| Ethnicity, n (%) |  |  |  |  |
| Hispanic or Latino | 62 (21.5) | 9 (12.7) | 13 (16.0) | 60 (17.8) |
| Not Hispanic or Latino | 216 (75.0) | 57 (80.3) | 63 (77.8) | 276 (81.9) |
| Unknown or Not Reported | 10 (3.5) | 5 (7.0) | 5 (6.2) | 1 (0.3) |
| BMI, median [Q1,Q3] | 28.5 [24.5,34.3] | 30.6 [24.6,36.0] | 27.3 [22.3,32.6] | 29.1 [24.1,35.9] |
| First intubation duration, median [Q1,Q3] | 5.0 [3.0,10.0] | 4.0 [3.0,10.0] | 6.0 [4.0,12.0] | 5.0 [2.0,11.0] |
| Unfavorable outcome, n (%) | 118 (41.0) | 38 (53.5) | 42 (51.9) | 143 (42.4) |

**Table 2.** Subset of feature descriptions across patient-ICU-days from the train, validate, test, and external datasets. Continuous variables are presented as median [Q1,Q3]. Categorical variables are presented as number (percentages). Full features are in Supplemental Table 2.

|  | Train | Validation | Test | External |
| --- | --- | --- | --- | --- |
| Number of days | 2264 | 594 | 831 | 2835 |
| ECMO flag, n (%) | 89 (3.9) | 34 (5.7) | 32 (3.9) | 168 (5.9) |
| CRRT flag, n (%) | 278 (12.3) | 58 (9.8) | 131 (15.8) | 226 (8.0) |
| PEEP, median [Q1,Q3] | 8.0 [5.0,12.0] | 7.5 [5.0,12.0] | 5.0 [5.0,10.0] | 8.0 [5.0,10.0] |
| FiO2, median [Q1,Q3] | 40.0 [30.0,50.0] | 40.0 [34.7,55.0] | 40.0 [30.0,55.0] | 45.0 [35.0,64.0] |
| Plateau Pressure, median [Q1,Q3] | 23.0 [18.0,28.0] | 26.0 [20.0,31.0] | 23.0 [18.0,29.0] | 23.0 [19.0,29.0] |

### Model results

The performances of the predictive models using different imputation strategies are summarized in Figure 2. The performance of all the models in predicting extubation was similar. An LSTM classifier using binned data achieved the best performance with a an area under the Receiver Operating Characteristic curve (AUROC) of 0.87 (95% CI 0.834-0.902) and an area under the Precision-Recall curve (AUPRC) of 0.382 (95% CI 0.269-0.492) on the test set (baseline rate of extubation/total days of 0.09, which would be the AUPRC for a no-skill model). Full metrics for all models before hyperparameter optimization are available in Supplemental Table 3 and full metrics for optimized models are available in Supplemental Table 4. In addition to the temporally split SCRIPT test set, we evaluated our top-performing model on data from an external hospital and found consistent performance across the two datasets (Figure 3).

**Figure 2.**
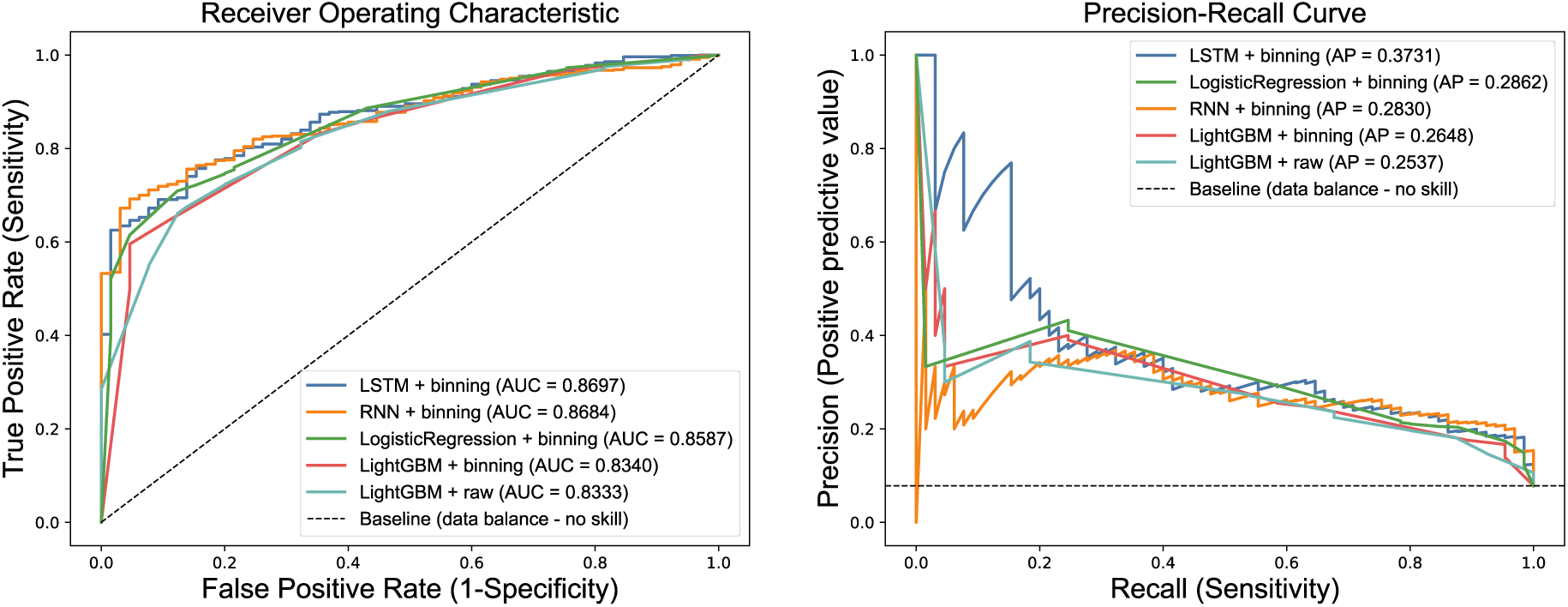
Performance Metrics of Different Machine Learning Models. The Receiver Operating Characteristic Curve (ROC), Precision-Recall Curve (PRC) plots of different machine learning models on same test set along with values of respective area under the curves, using each model’s best-performing imputation method, including Extreme Gradient Boosting (XGBoost), Recurrent Neural Network (RNN), and Long Short-Term Memory (LSTM), on the test set, using different imputation strategies (raw and binning, detailed in Methods). Curves displayed are for a single pass through the test set. Full metrics and confidence intervals for the top optimized models shown are in Supplemental Table 4.

**Figure 3.**
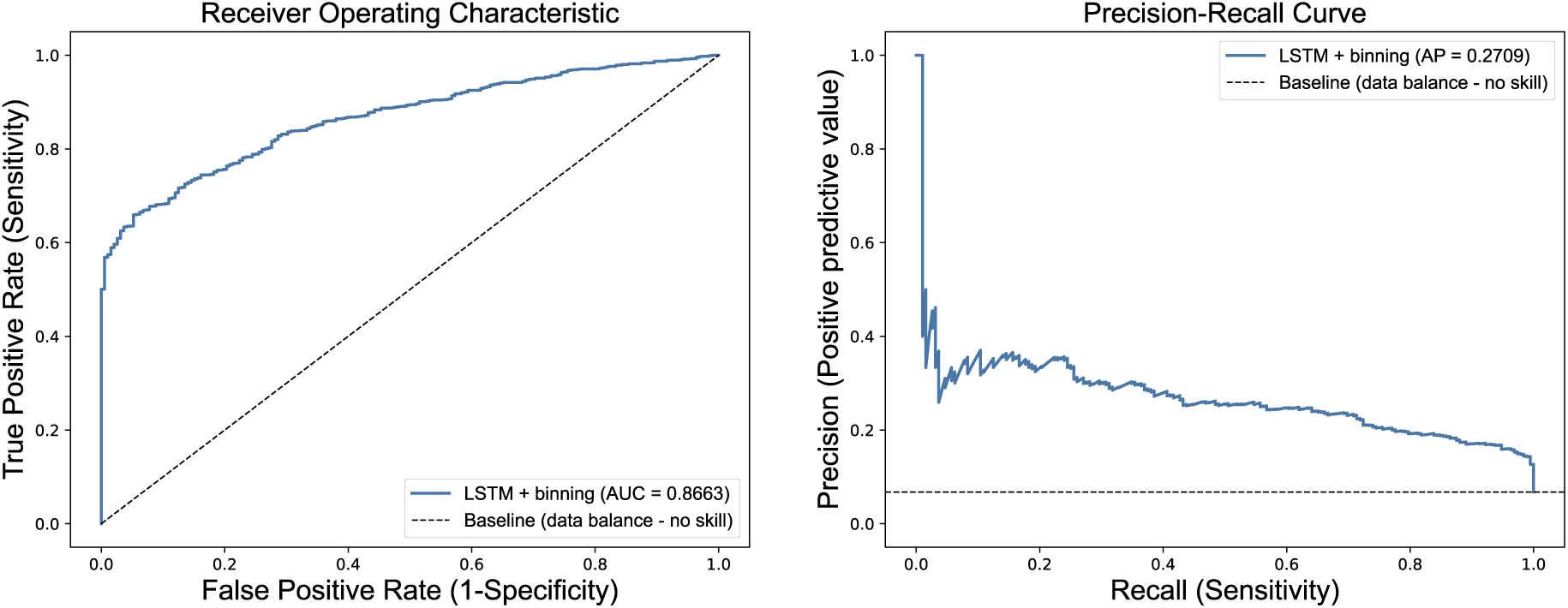
Model performance on external test cohort. We applied our top-performing binned LSTM model (based on AUROC) to a patient cohort from a different hospital system as an external test. **(A)** ROC and **(B)** PRC curves show similar performance to the SCRIPT test set in Figure 2. Performance on this external dataset (0.87 AUROC [95% CI 0.848-0.885], 0.274 AUPRC [95% CI 0.222-0.332]) is consistent with performance on the internal test set (0.87 AUROC [95% CI 0.834-0.902], 0.3782AUPRC [95% CI 0.269-0.492]).

### Imputation/encoding strategies affect model performance

We compared the performance of different imputation strategies across models as well. The best encoding strategy was binning, which encodes data presence or missingness, and then bins continuous variables into quartiles. The next was using raw data without any imputation, in models that could accommodate this such as XGBoost and LightGBM. Simpler but commonly employed strategies such as mean imputation performed consistently the worst across models all models (Supplemental Table 3).

### Feature importance

The top ten features contributing to the predictive performance of the XGBoost model were identified using SHAP (SHapley Additive exPlanations) values (Figure 4). They include markers of the patient’s mental status, as documented by Glascow Coma Scale (GCS), as well as markers of the patient’s lung physiology, as documented by presence of ventilator measurements such as plateau pressure. Similar features were present on LSTM ablation feature importance analysis, with top feature importance including RASS score, respiratory rate, plateau pressure, and GCS motor response. RASS score, plateau pressure, and GCS motor response are also represented by different binned features in the top ten feature coefficients for Logistic Regression.

**Figure 4.**
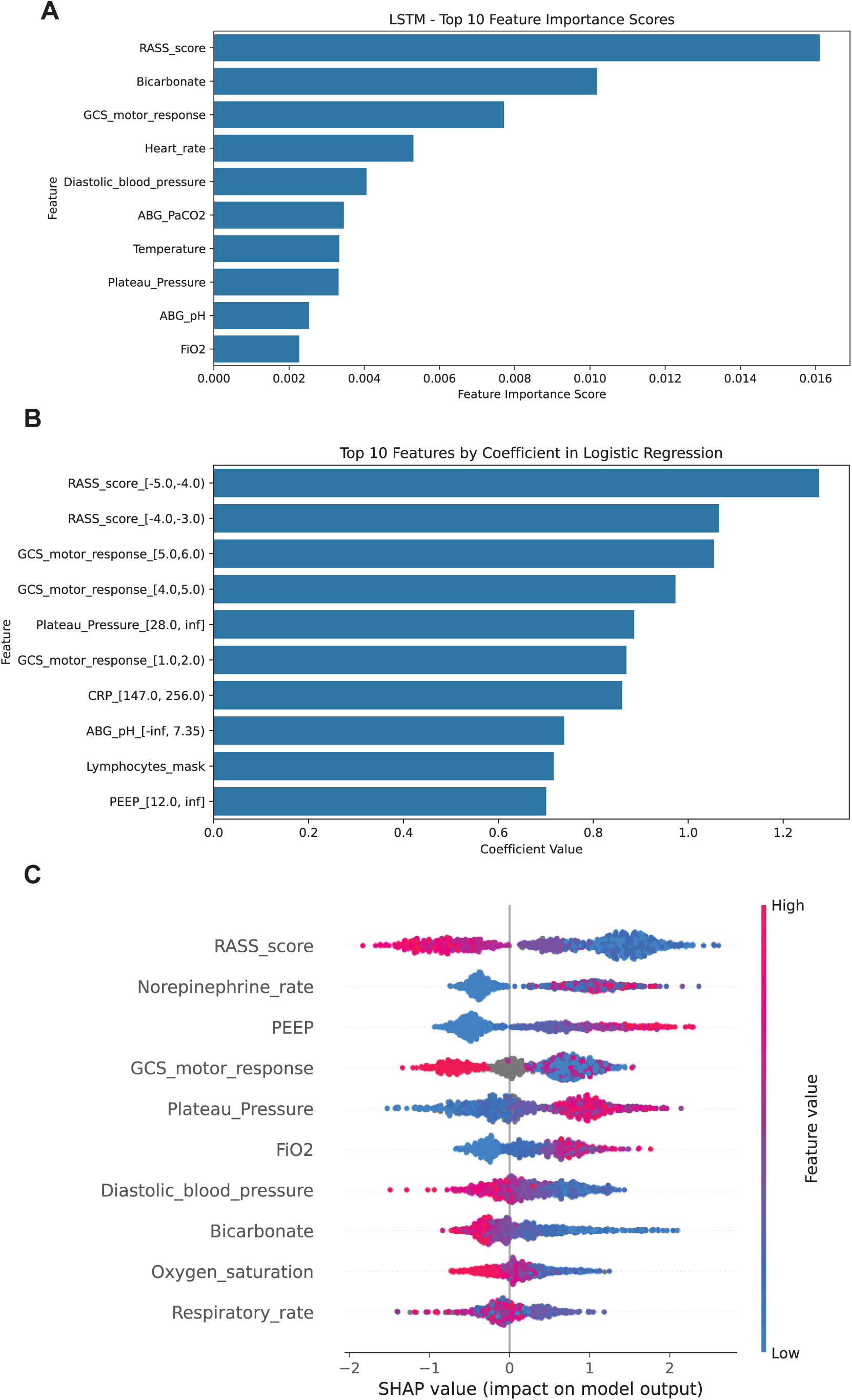
Feature importance plots. **(A) Feature importance ablation plots** for an LSTM model predicting next-day successful extubation provide insights in the significance of each feature. In these plots, each feature is a row, and the x axis represents how important that feature is. This feature importance is calculated by doing 37 iterations (number of features) while masking an individual feature each time and seeing the decline in AUC on the test set without that feature available. **(B) Logistic regression coefficients** for each feature when applied to binned data. Bins for Rass score, GCS motor response, and plateau pressure show concordance with feature importance in (A) and (C). **C) SHAP (Shapley Additive Explanations) plots** for an XGBoost model predicting next-day successful extubation. In these plots, each feature is a row, with the color indicating the feature’s value (e.g., red for higher values and blue for lower values). The position of the bar on the plot represents the feature’s Shapley value, with features on the right side contributing positively to the prediction of staying intubated and those on the left side contributing to getting extubated. Thus, the length and direction of the bars illustrate the strength and direction of influence that each feature has on the model’s decisions. For example, a high plateau pressure (red) is associated with increased likelihood of remaining intubated (right-ward of zero).

### Missed opportunities and preventing failed extubations

Our model often predicted patients could be considered for extubation in the days preceding their actual extubation, suggesting opportunities to consider earlier extubation (Figure 5A). This is a stringent way of evaluating the model as only the earliest extubation prediction is used for analysis (Supplemental Methods). We also tested on failed extubations (requiring reintubation within two days) not seen by the model during training. Our best model predicted extubation failure in 35.4% (17/48) of these cases.

**Figure 5.**
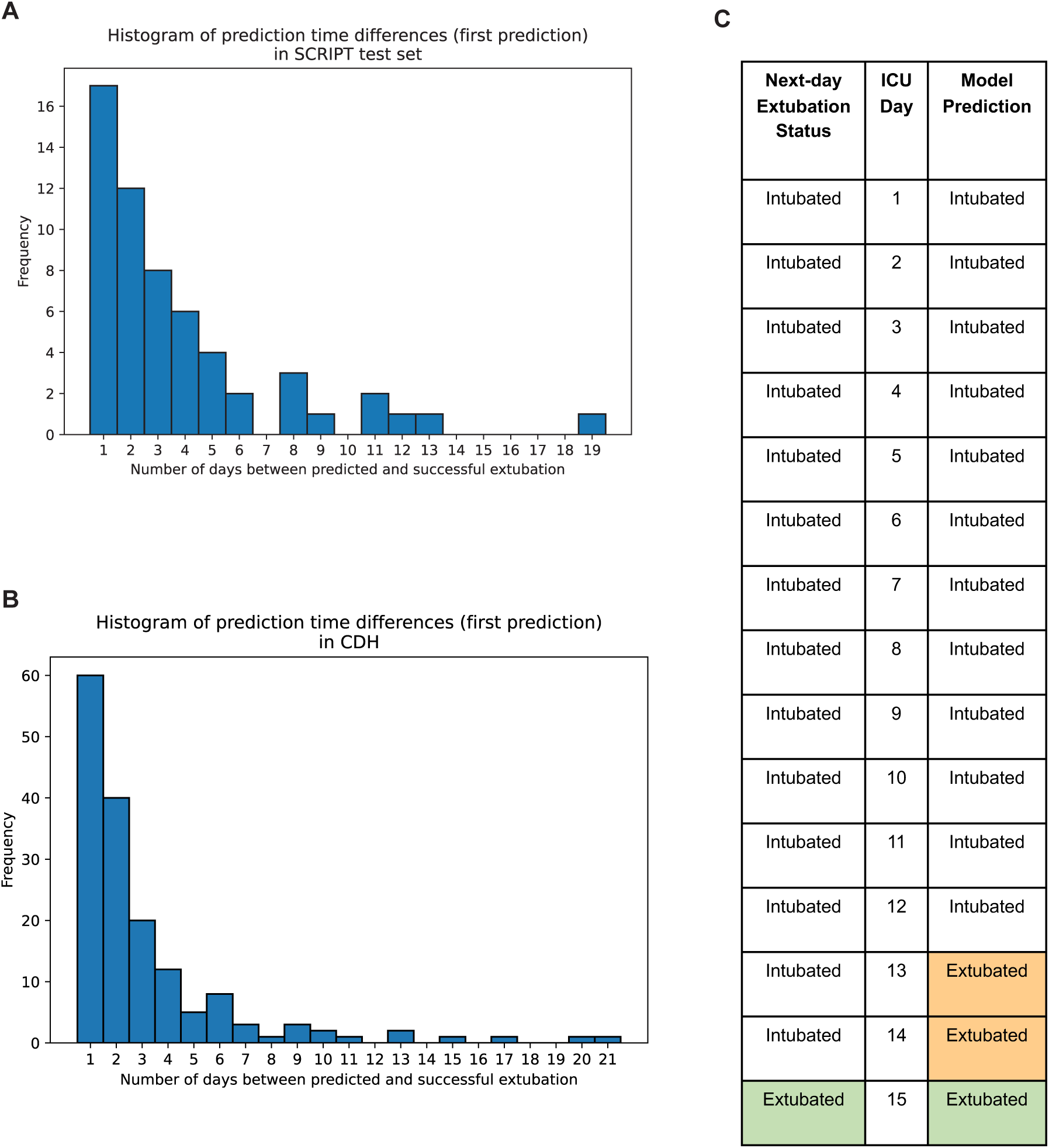
Examining discrepancies between model prediction and time of extubation. **(A)** For each intubation sequence preceding a successful extubation in the SCRIPT test set, we examined the first instance predicting next-day extubation (Supplemental Methods). Many model predictions, if not exactly one day before extubation as intended (29.3%), were within two days (50%) or three days (63.8%). **(B)** The same analysis as (A) performed on the external test set, where we report 37.3% of first next-day extubation predictions occurring within one day of successful extubation, 62.1% within two days, and 74.5% within three days. **(C)** These early extubation predictions may be missed opportunities where a clinical team could have potentially extubated the patient earlier. In the example, the model first predicted extubation 3 days prior to the true successful extubation.

## Discussion

Identifying patients who are ready for extubation remains a challenging clinical problem in the ICU. Although ventilator-weaning protocols have been shown to outperform routine physician-driven care,^4,25^ the adoption and standardization of these practices varies.^5,26,27^ Barriers to the adoption of some protocols include their requirement for trained personnel^28^ and in some cases specialized equipment.^29^ We built several machine learning models that incorporated features included in the electronic health record in most ICU settings. Several of these models performed well with the best model providing an AUROC above 0.85. These analyses also suggest that HER-based classifiers would have identified some patients who might have been considered for earlier SBTs and extubation. We found 63.8% of predicted extubations in cases of successful extubation occurred within three days of actual extubation, suggesting potential earlier extubation could have been possible if the team were following model-driven care. We also found, when looking at sequences where a patient had an unsuccessful extubation, our model would have recommended against extubation in 35.4% of cases.

The models we used allow us to identify clinical features that drive their predictions. Protocol driven weaning which has been rigorously shown to shorten the duration of mechanical ventilation usually includes an assessment of mental status, oxygenation, hemodynamic stability, an absence of ongoing infection, and physiologic measures of lung mechanics.^7,30^ Our models were informed by clinical features that reflect these assessments, lending clinical plausibility to their predictions. Furthermore, models that treated each day independently performed similarly or just slightly inferior to sequential models that used information from multiple days, consistent with efficacy of protocols that assess readiness for spontaneous breathing on a daily basis irrespective of clinical context.^3^

We also examined the effect of different imputation strategies on model performance. Commonly used strategies, such as filling in missing values with the data’s mean value, performed significantly worse than maintaining missingness using either encoding strategies such as binning or working with models such as XGBoost that can inherently handle missing data. This strategy can be particularly useful for complex models like deep learning algorithms where data in a medical context can have such extreme variance.

SCRIPT is a highly curated dataset, with a panel of five critical care physicians meeting weekly to review each admitted patient and their outcomes.^14^ A full-time research team collects enrollment and end of study data, including if a patient was transitioned to comfort-focused care. Since this is not possible for larger scale datasets, we compiled a set of pragmatic rules based on our experience (Supplemental Methods). We performed significant manual curation of our dataset of over 200 patient charts, confirming manually on the front end EHR unusual cases of intubation, which can be difficult to parse out using only flowsheet data.^31^ We believe that our careful data cleaning and manual adjudication processes were key in developing a clinically important model using features previously known to be clinically important to inform model development.^32^ Compared with other machine learning models to predict extubation^33^ or shift off full ventilatory support,^34^ our work obtains good predictive performance when applied to a population of critically ill patients in both an internal test dataset and when applied to a distinct medical ICU setting. Ensuring a sufficient time gap between data gathering and the prediction of interest is a crucial step in mitigating the risk of data leakage in a machine learning model. In contrast to studies that make predictions on the next time period without any gap for intervention,^35^ we specifically use only information from midnight to 8AM rather than the entire day so the model can offer this information to the multidisciplinary team on morning rounds to inform clinical decision-making.

### Limitations

First, our model was trained using clinically adjudicated data from a quaternary care referral health system that cares for severely ill patients, reflected in the substantial mortality and long duration of mechanical ventilation, which might limit its relevance in other ICU settings. Nevertheless, the model performed well in a medical ICU from a community-based hospital, suggesting carefully curated data from relatively small populations can be used to train models for generalized use. Importantly, however, the community-based hospital ICU we used for validation uses the same EHR as our academic medical center. We do not know if different health record instances would affect performance and this remains to be evaluated in future work. Second, we limited the information used to train our models to physiologic factors that are measured in most ICUs. This necessarily excluded factors, such as continued intubation for a procedure, that might have influenced clinicians’ decision on readiness for extubation. Our dataset is small compared with larger ICU cohorts such as MIMIC or eICU^36,37^ and future work validated on these datasets is needed. Our study focused on MICU patients with suspected pneumonia, so generalizing these findings to other ICU settings, for example surgical or cardiac ICUs, will require further work. While our model supports the feasibility of a machine learning model to identify patients who might be considered for extubation, we have not yet deployed it in a clinical setting. Nevertheless, we designed our predictor with an eye towards deployment, only using data from midnight to 8AM. In the future, we envision providing model output to a clinical team on rounds as a Bayesian pre-SBT probability of extubation success. Alternatively, the model could be incorporated into the EHR to automatically trigger a clinical decision support alert to bedside nurses or respiratory therapists suggesting initiation of a SBT to help optimize the timing of extubation, consistent with best evidence.^4^ We hope that this can help both shorten intubation duration and prevent failed extubations, to improve the care of critically ill patients.

## Supporting information

Tripod Checklist

NU SCRIPT Study Investigator List

## Acknowledgements

BioRender was used for figure generation.

## Human Ethics and Consent to Participate

This study was approved by the Northwestern University Institutional Review Board with study IDs STU00204868 and STU00216678.

## Data availability

A significant portion of this data has been already made available through PhysioNet at https://physionet.org/content/script-carpediem-dataset/1.1.0/, a future update will include new patients and updated data since the publication of the original dataset. Code for processing and analysis are available at https://github.com/NUPulmonary/2024_Fenske_Peltekian.

## Author contributions

Conceptualization: CAG, AA, AVM, GRSB, RGW

Methodology: SF, AP, AA, CAG

Data acquisition: MK, AP

Programming/analysis/visualization: SF, AP, NSM, AA, CAG

Chart review: MZ, KG, MB, CAG

Drafting: SF, AP, AA, CAG

Editing: all authors

## Supplemental methods and materials for *Developing and validating machine learning model to predict successful next-day extubation in the ICU*

### Cohort information

Our study involved patients who had received mechanical ventilation and were admitted to an intensive care unit (ICU) as part of the Successful Clinical Response in Pneumonia Therapy (SCRIPT) study conducted at Northwestern Memorial Hospital (NMH) between June 2018 and April 2023. Patients who had multiple ICU stays during the same hospitalization had their stays numbered consecutively but did not contribute data between their ICU stays. Multiple hospitalizations of the same patient are reported as separate patients, as SCRIPT enrollments are unique to each hospitalization. For patients who had multiple sequences of intubation and extubation in a given ICU stay, we used only the first intubation/extubation sequence.

### EHR data

Electronic health record (EHR) data are compiled by the Northwestern Medicine Enterprise Data Warehouse (EDW), the primary data repository for clinical data at Northwestern Medicine. Approximately 150 data sources (including the main EHR system, Epic) are loaded into the EDW on a nightly basis. The data are primarily loaded using Microsoft technologies (Visual Studio, SSIS, etc.) and scheduled to load via the SQL Server Job Agent. Data engineers and architects on the EDW team then combine the data sources using custom SQL scripts, Visual Studio, SSIS, etc. to create datamarts. Analysts on the EDW team then work with the datamarts to create reports, dashboards, and extracts validated with clinician input. Over two hundred charts were manually reviewed by physicians to ensure data pull fidelity. Questions were brought to a committee meeting comprised of five critical care physicians and consensus was obtained before moving forward. Each patient chart was reviewed by the research study team on study close to gather information including transition to comfort measures only.

### Clinical parameters

We selected 37 clinical features considered representative of those that physicians would consider during daily ICU rounds. We compiled the status of intubation, extracorporeal membrane oxygenation (ECMO), renal replacement therapy (hemodialysis [HD], and continuous renal replacement therapy [CRRT]), sedation parameters (Glasgow Coma Scale [GCS] subscores of eye opening, motor response, verbal response, and Richmond Agitation Sedation Scale [RASS]), lung injury (arterial pH, PaO2, PaCO2, PEEP, FiO2, plateau pressure, lung compliance, minute ventilation, and oxygen saturation), hemodynamics (norepinephrine rate in mcg/kg/min, mean arterial pressure, systolic and diastolic blood pressure, lactic acid, hemoglobin, and bicarbonate), renal (creatinine, and aforementioned HD and CRRT flags), inflammatory markers (WBC count, neutrophil count, platelets, procalcitonin, C-reactive protein, D-dimer, lactate dehydrogenase, ferritin, bilirubin, albumin, and lymphocytes), vital signs (temperature, heart rate, respiratory rate). When multiple measurements were available for the same time block, they were aggregated by mean. Outliers were removed prior to aggregation by using predefined ranges for each measurement. Full details are available in our code at https://github.com/NUPulmonary/2024_Fenske_Peltekian.

### Time blocking

Since the majority of extubations occur after 8AM (Supplemental Figure 1), we aggregated our dataset using only information from midnight to 8AM. This strategy would be optimal for future deployment to allow information and model predictions to be presented to the clinical team during rounds.

### Annotating flags

ECMO_flag was labeled as 1 if that day lay on or in between the first and last days of recorded ECMO. Hemodialysis_flag was labeled as 1 if the patient was ever on Hemodialysis between midnight and 8AM that day. CRRT_flag was also labeled as 1 if the patient was ever on CRRT between midnight and 8AM that day.

### Continuous features

Features measured on a continuous numerical scale were aggregated by mean from all recordings between midnight and 8AM that day.

### Categorical features

RASS and GCS scores were the only ordinal features in the dataset. Because they are still a numerical scale, they were aggregated the same way continuous features were.

### Annotating extubation status

To label days as intubated or extubated, we parsed through the EDW reports of oxygen support devices (ranging from nasal cannula to invasive mechanical ventilation) and employ a series of rules to determine the extubation status of each day. Full details are present in our code repository at https://github.com/NUPulmonary/2024_Fenske_Peltekian. In most cases, a patient is maintained entirely as intubated or extubated between midnight to 8AM. Extubation status was labeled according to data from the oxygen device report for that day.

When pulling data from the EDW, days were marked as intubated based on the presence of at least two instances of intubation or if one of the following was documented: PEEP, plateau pressure, static compliance, or minute ventilation. The cutoff of 8AM was purposely chosen to obtain the cleanest data such that transitions on/off the ventilator do not confound that extubation status of a patient day. There are two other broad classes: a patient being intubated/extubated before 8AM. If a patient is intubated before 8AM, this day is excluded from modeling due to a mix of intubated and extubated data, and the following day, assuming they remain intubated from midnight to 8AM, would mark the first ICU day presented to the model. If a patient is extubated before 8AM, the day is marked as extubated. Box 1 provides a summary of our inclusion criteria for training a model on successful next-day extubation. Box 2 outlines further manual checks that were performed to ensure proper labeling of extubation status between 12AM and 8AM on each day.

#### Box 1.

##### Preserving only successful extubation cases.

- Exclude patient-days with tracheostomy, as ventilator liberation in this population is different and lower stakes than patients with endotracheal tubes.
- Excluding cases where reintubation is required within two days of extubation.
- Excluding cases of extubation while the patient remains on ECMO.
- Excluding cases of palliative extubation. In our dataset, this was gathered by the research team and CAG performed additional manual review. A pragmatic way of labeling this in the absence of chart review would be to exclude cases where a patient died within 48 hours of extubation.

#### Box 2.

##### Using O2 device report to assign extubation status.

1. If documented extubation event on an intubated day, assign as extubated.
2. If extubated day has at least three or is entirely documentations of intubation, label as intubated.
3. If patient is re-intubated within two days of an extubated day, label the extubated day(s) as failed extubation.
4. If within the same day, a patient is intubated, then extubated, then has at least three documentations of intubation following the extubation event or the next day is intubated, label as failed extubation. Here we require a minimum of three documentations of intubation following the extubation event due to one-off ventilator reads that will show up during the extubation documentation. If the next day is intubated, it is safe to assume that the intubation documentation is correct.
5. If patient was extubated on their last day in the ICU and we have documentation that it was a pallative extubation, then label the preceding extubated days as CMO.
6. If patient was extubated on their last day in the ICU and die within 48 hours of the day of extubation, label the day(s) as CMO.
7. Flag all days that have at least three documentations of trach collar. These patients will be withheld from analysis.
8. Label all extubated days preceding intubation during a given ICU stay as preintubated.

### Multiple intubation sequences

Sequences including failed extubation are excluded from training. Cases of extubation while still on ECMO and preintubation days are also excluded from training. Cases with one ICU day are naturally excluded as there is no next-day extubation status to predict. We further consolidated our training dataset by excluding sequences of reintubation.

### All-inclusive model

We initially tried an all-inclusive model that included all days. This included pre-intubation, CMO, failed, and successful extubation days. This was easier to assess and presented a better general purpose model design, but the model failed to capture the differences between intubated and extubated days. This was due to the noise introduced by CMO, failed, and pre-intubation days that made it difficult for the model to learn when a patient was ready for extubation. Designing a model with the more focused goal of predicting *successful* extubation, we consolidated our time series sequences and were able to see the model learn the differences between patient-days on which a successful versus unsuccessful extubation would be predicted to occur.

### Imputation Strategies

- No imputation, called ‘raw’: as some models such as XGBoost can inherently handle missing values, no imputation was performed in ‘raw’ models.
- Mean Imputation: The missing values were initially replaced with the mean of the respective feature derived from the training data. This mean value was then applied universally to any absent data points in both training and test sets.
- Building upon the work of Fiddle,^17^ a modified version was employed as an encoding strategy. Continuous features were binned into quartiles derived from the training set, subsequently encoding each as either 1 or 0 based on its bin membership. All absent features were encoded using a fifth feature, represented by a 1, which signifies a missing value. Thus, for a given feature, only exactly one of the five derived features would be encoded as 1, and the other four encoded as 0. For example, if someone had a temperature of 97.1, the value would be encoded as a 1 in the lowest bin of temperature quartiles, with the rest of the bins being zero (Bin 1: 1, Bin 2: 0, Bin 3: 0, Bin 4: 0, Mask Bin: 0). If their temperature value is missing for that day, it would be encoded in the mask bin. Quartiles, or the boundaries for each bin, were established based on the range of values in the train set, and the test set data is placed into these same bins.

### Deep learning and boosting models in sequential data prediction

#### Deep Learning Architectures

RNN and LSTM

In deep learning, Recurrent Neural Networks (RNNs) and Long Short-Term Memory networks (LSTMs) are valuable for sequential data prediction. Our models consisted of an input layer designed to handle sequences of varying lengths, padded with zeros. We included a masking layer so that these padded values do not interfere with the model’s computations by skipping time steps with zero input. RNNs can handle data in a sequential manner, capturing time-step patterns and remembering historical data. However, they can experience issues with the vanishing gradient problem and can propagate errors across predictions. LSTMs address the vanishing gradient issue, handling long sequences with their advanced memory capabilities. They do also share the problem of error propagation and are computationally more complex than standard RNNs.

### Padding Sequences

For the deep learning models, sequences of varying lengths were encountered. To make them consistent in length, they were padded with 0s. This ensures that the model receives input of a uniform shape, a requirement for efficient batch processing.

### Boosting Models

Boosting models, such as XGBoost and LightGBM, treat each data point independently. They are calibrated using isotonic regression for better alignment with the true data distribution, which is often used in imbalanced datasets. Boosting models are efficient against missing data and do not let incorrect predictions influence subsequent ones since they handle prediction problems non-sequentially. They often outperform other traditional machine learning algorithms but lack the sequential data understanding and may overfit if not properly tuned. Logistic regression also treats data points independently but cannot handle missing data.

### Hyperparameter Optimization

Hyperparameter tuning was performed on the top five best performing models, with details of specific hyperparameters tried in Supplemental Table 5.

### Key Metric Observations

#### AUROC (Area Under the Receiver Operating Characteristic Curve)

Measures the ability of a model to distinguish between classes. It is represented as a curve plotting the true positive rate against the false positive rate at various threshold settings.

#### AUPRC (Area Under the Precision-Recall Curve)

Plots precision (the ratio of true positives to the sum of true and false positives) against recall (the ratio of true positives to the sum of true positives and false negatives). This curve provides a trade-off between precision and recall across different thresholds, making it useful for evaluating models where one class is significantly underrepresented.

#### Accuracy

Ratio of correctly predicted observations to the total observations. It represents the proportion of true results (both true positives and true negatives) among the total number of cases examined.

#### F1 Score

Provides a balance between precision and recall. The F1 Score reaches its best value at 1 (perfect precision and recall) and worst at 0.

#### Recall

Recall (sensitivity) measures the proportion of actual positives correctly identified by the model. It is calculated as the ratio of true positives to the sum of true positives and false negatives.

#### Precision

Measures the proportion of positive predictions that are correct. It is calculated as the ratio of true positives to the sum of true positives and false positives.

### Confidence intervals

Confidence intervals for general performance metrics were generated by bootstrapping. Bootstrapping was done through 1000 passes through the dataset where each pass takes n days from the dataset with replacement, where n is the number of days in the test set. The inner 95^th^ percentile of each metric is the reported 95% CI.

### Threshold for binary predictions

Set using the class imbalance in the train set. Specifically, the ratio of next-day intubated days to all days (0.91).

### Feature Importance

#### SHAP values

SHAP values are a useful tool for explaining the output of any machine learning model. They measure the impact of each feature on a particular prediction in comparison to the model’s baseline output. For our boosting models, SHAP (using the summary_plot method) values give us the marginal contribution of each feature towards the prediction for every individual instance. This explains importance features, and also how a specific feature can influence a particular prediction.

#### LR coefficients

For Logistic Regression models, each feature contains a coefficient where greater coefficients imply greater effect on model output.

#### Ablation

For RNN and LSTM models, an ablation procedure was performed to quantify a feature’s impact on model performance. For each feature in the test set, all values were converted to NA, and the change in AUROC from such data from the original data was reported as the feature importance score. For example, when assessing the feature importance of RASS score in a binned LSTM model, each RASS score value would shift from a 1 in whichever bin it fell in to a 1 in the RASS_score_mask column. Based on Figure 4, we know this led to a decrease in AUROC, implying RASS score is an important feature to the model.

### Evaluating early model predictions

To investigate the timing of extubation predictions, we investigated where incorrect model predictions occurred in reference to true extubation events. To provide an unbiased quantification of this, we subset on all ICU stays containing a successful extubation, and examined where the model first predicted extubation during these ICU stays. We report that a significant amount of incorrect model predictions are within three days of the true extubation event (Figure 5).

#### Supplemental Figures and Tables

**Supplemental Figure 1:**
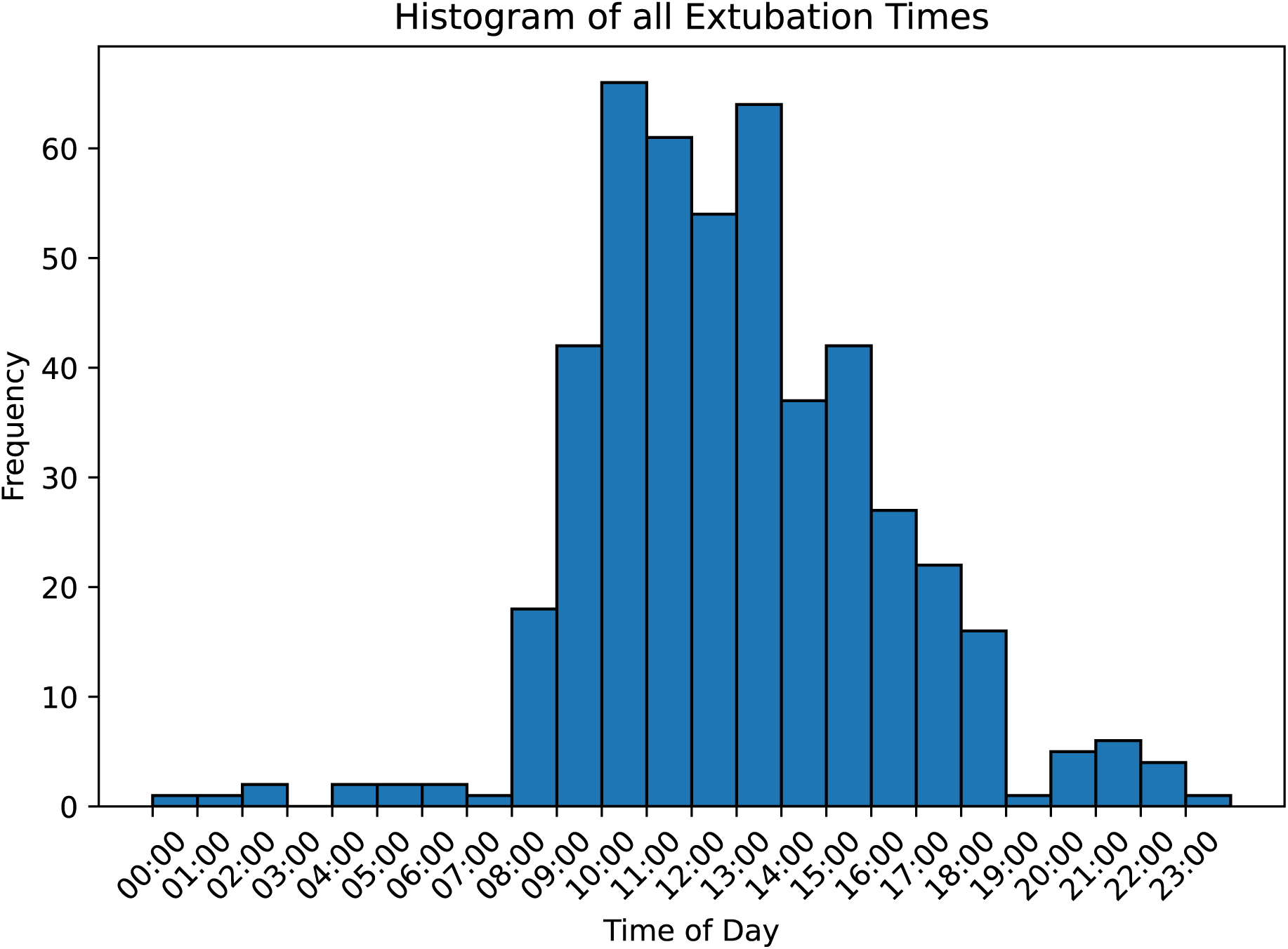
Histogram of extubation times. Most extubation events occur between 8am and 6pm, with just 2.3% occurring before 8am. This informed our choice to create a model that provides a daily prediction at 8am based on the most recent data from that day.

**Supplemental Figure 2:**
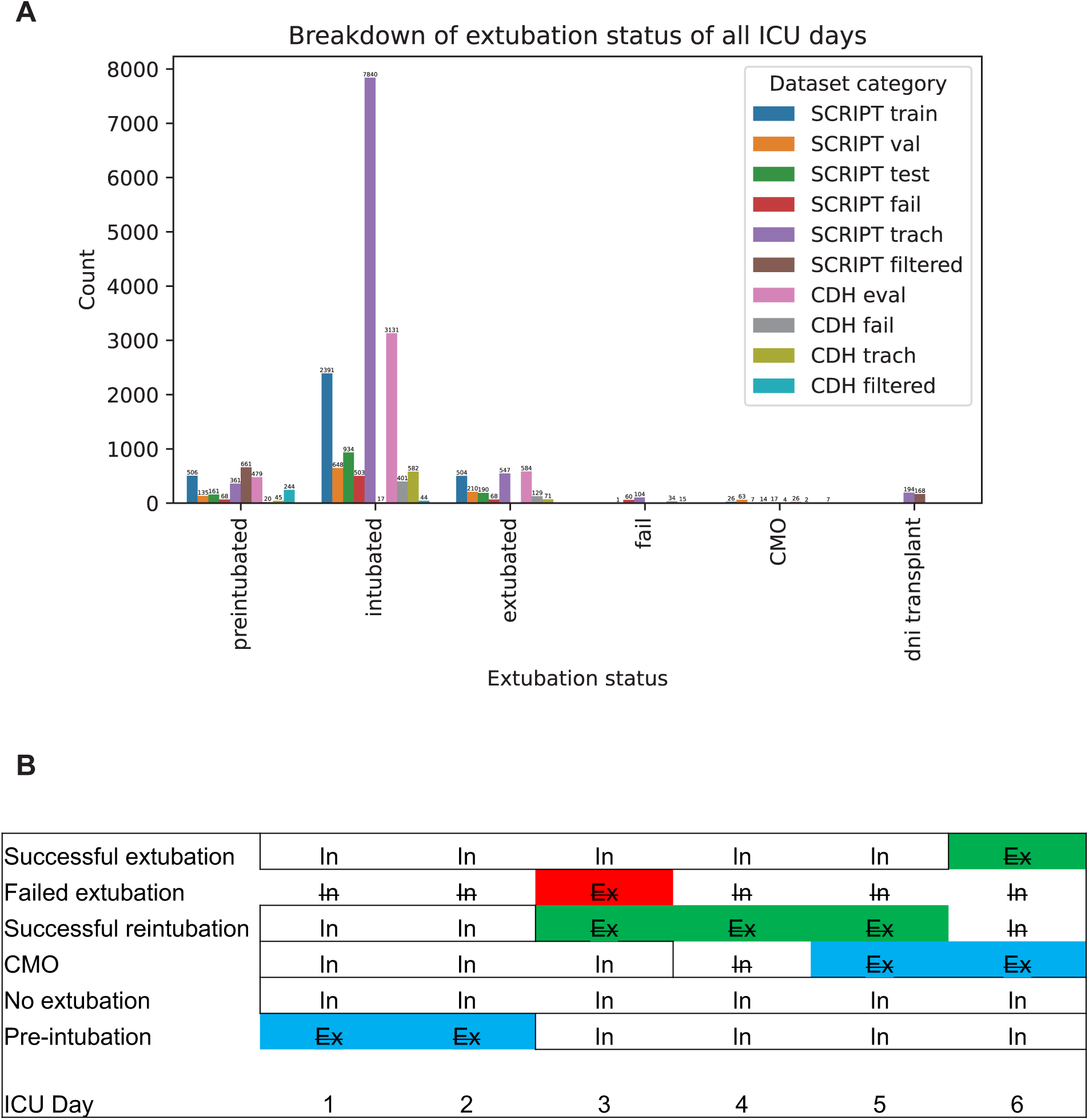
Types of Intubation/Extubation Sequences. This figure categorizes various sequences of intubation and extubation events encountered in clinical practice. It illustrates distinct patterns such as planned extubations following successful weaning without need for reintubation, failed extubation, and palliative extubation when moving to comfort care. Understanding and classifying these sequences is crucial for optimizing model training. **(A)** number of days for each type of intubation/extubation day. Tran/val/test represent patient days present in each split. Fail represents days held out as a separate test set due to a failed extubation event. Filtered represents all days excluded from model training and evaluation. **(B)** Inclusion criteria for modeling successful next-day extubation. Sequences of failed extubation, preintubated days, CMO and the days before, and reintubation sequences are excluded from the train/test set.

**Supplemental Table 1.**
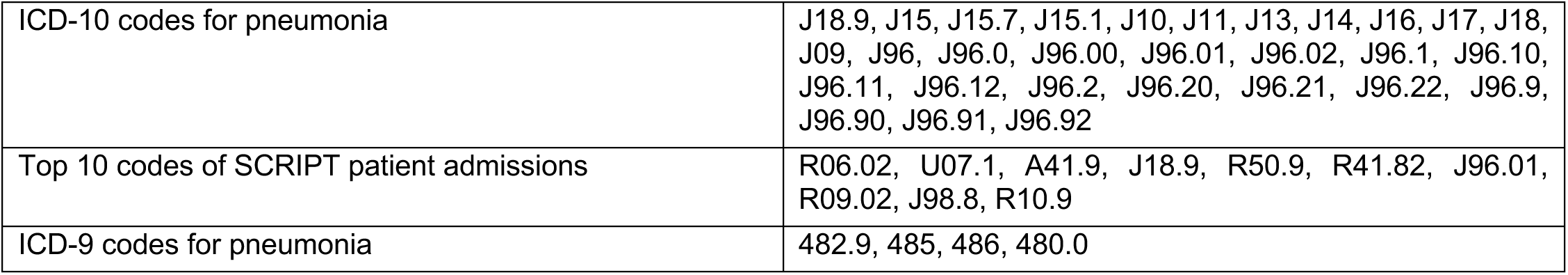
ICD diagnosis codes used for pulling patient data for the CDH cohort.

|  |  |
| --- | --- |
| ICD-10 codes for pneumonia | J18.9, J15, J15.7, J15.1, J10, J11, J13, J14, J16, J17, J18, J09, J96, J96.0, J96.00, J96.01, J96.02, J96.1, J96.10, J96.11, J96.12, J96.2, J96.20, J96.21, J96.22, J96.9, J96.90, J96.91, J96.92 |
| Top 10 codes of SCRIPT patient admissions | R06.02, U07.1, A41.9, J18.9, R50.9, R41.82, J96.01, R09.02, J98.8, R10.9 |
| ICD-9 codes for pneumonia | 482.9, 485, 486, 480.0 |

**Supplemental Table 2.**
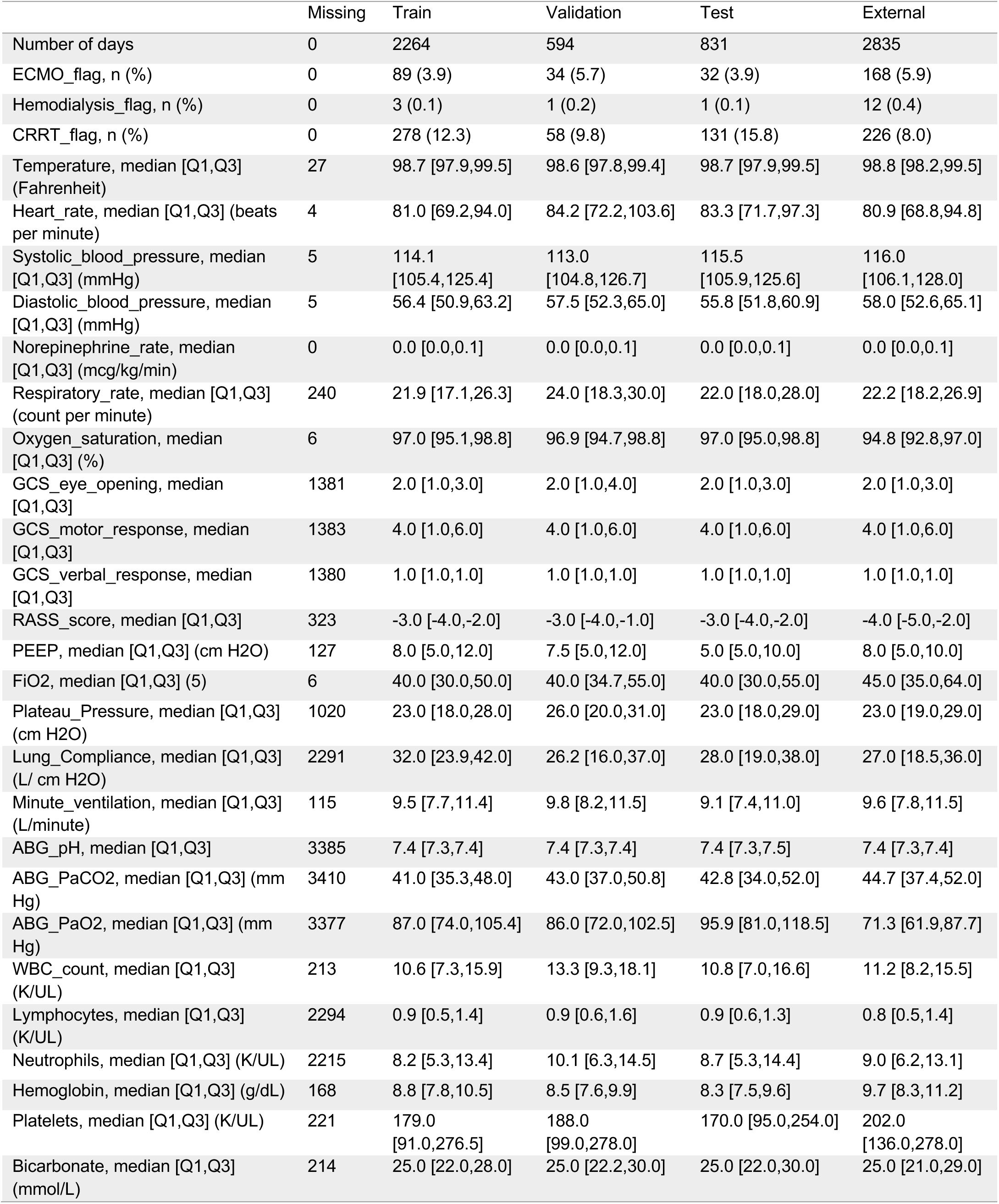
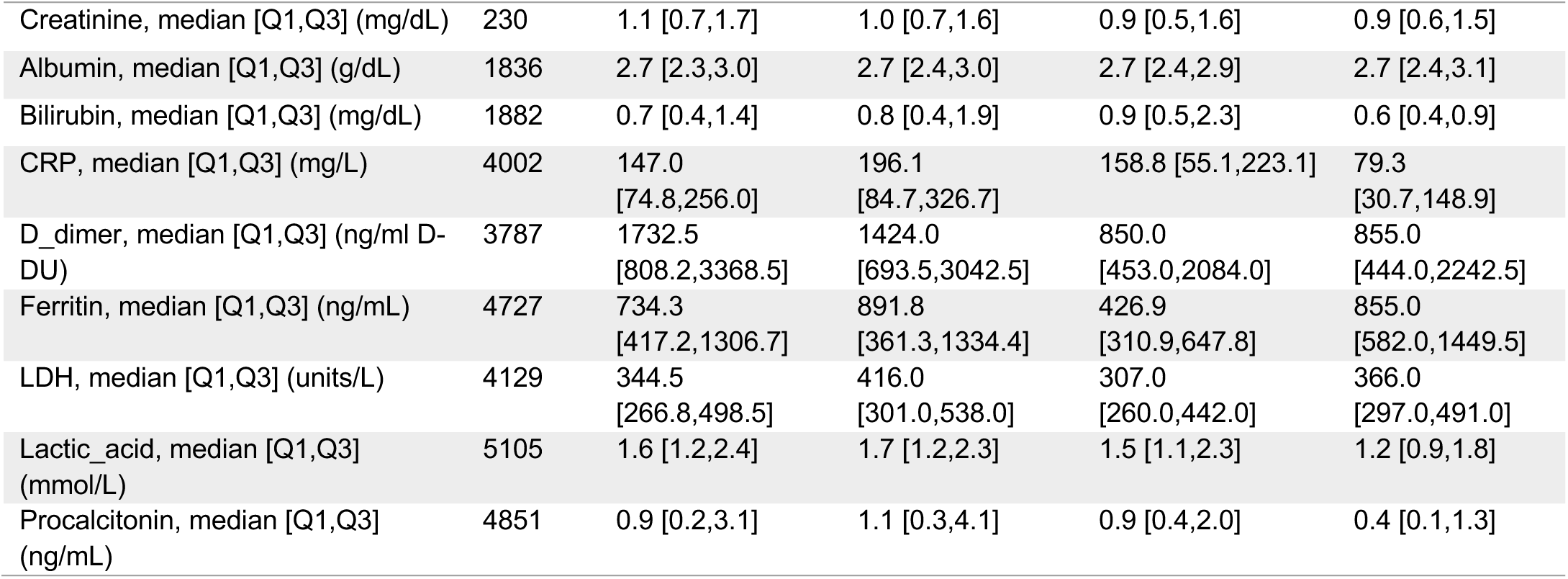
Full list of features across patient-ICU-days from the train, validation, test, and external datasets.

|  | Missing | Train | Validation | Test | External |
| --- | --- | --- | --- | --- | --- |
| Number of days | 0 | 2264 | 594 | 831 | 2835 |
| ECMO_flag, n (%) | 0 | 89 (3.9) | 34 (5.7) | 32 (3.9) | 168 (5.9) |
| Hemodialysis_flag, n (%) | 0 | 3 (0.1) | 1 (0.2) | 1 (0.1) | 12 (0.4) |
| CRRT_flag, n (%) | 0 | 278 (12.3) | 58 (9.8) | 131 (15.8) | 226 (8.0) |
| Temperature, median [Q1,Q3] (Fahrenheit) | 27 | 98.7 [97.9,99.5] | 98.6 [97.8,99.4] | 98.7 [97.9,99.5] | 98.8 [98.2,99.5] |
| Heart_rate, median [Q1,Q3] (beats per minute) | 4 | 81.0 [69.2,94.0] | 84.2 [72.2,103.6] | 83.3 [71.7,97.3] | 80.9 [68.8,94.8] |
| Systolic_blood_pressure, median [Q1,Q3] (mmHg) | 5 | 114.1 [105.4,125.4] | 113.0 [104.8,126.7] | 115.5 [105.9,125.6] | 116.0 [106.1,128.0] |
| Diastolic_blood_pressure, median [Q1,Q3] (mmHg) | 5 | 56.4 [50.9,63.2] | 57.5 [52.3,65.0] | 55.8 [51.8,60.9] | 58.0 [52.6,65.1] |
| Norepinephrine_rate, median [Q1,Q3] (mcg/kg/min) | 0 | 0.0 [0.0,0.1] | 0.0 [0.0,0.1] | 0.0 [0.0,0.1] | 0.0 [0.0,0.1] |
| Respiratory_rate, median [Q1,Q3] (count per minute) | 240 | 21.9 [17.1,26.3] | 24.0 [18.3,30.0] | 22.0 [18.0,28.0] | 22.2 [18.2,26.9] |
| Oxygen_saturation, median [Q1,Q3] (%) | 6 | 97.0 [95.1,98.8] | 96.9 [94.7,98.8] | 97.0 [95.0,98.8] | 94.8 [92.8,97.0] |
| GCS_eye_opening, median [Q1,Q3] | 1381 | 2.0 [1.0,3.0] | 2.0 [1.0,4.0] | 2.0 [1.0,3.0] | 2.0 [1.0,3.0] |
| GCS_motor_response, median [Q1,Q3] | 1383 | 4.0 [1.0,6.0] | 4.0 [1.0,6.0] | 4.0 [1.0,6.0] | 4.0 [1.0,6.0] |
| GCS_verbal_response, median [Q1,Q3] | 1380 | 1.0 [1.0,1.0] | 1.0 [1.0,1.0] | 1.0 [1.0,1.0] | 1.0 [1.0,1.0] |
| RASS_score, median [Q1,Q3] | 323 | -3.0 [-4.0,-2.0] | -3.0 [-4.0,-1.0] | -3.0 [-4.0,-2.0] | -4.0 [-5.0,-2.0] |
| PEEP, median [Q1,Q3] (cm H2O) | 127 | 8.0 [5.0,12.0] | 7.5 [5.0,12.0] | 5.0 [5.0,10.0] | 8.0 [5.0,10.0] |
| FiO2, median [Q1,Q3] (5) | 6 | 40.0 [30.0,50.0] | 40.0 [34.7,55.0] | 40.0 [30.0,55.0] | 45.0 [35.0,64.0] |
| Plateau_Pressure, median [Q1,Q3] (cm H2O) | 1020 | 23.0 [18.0,28.0] | 26.0 [20.0,31.0] | 23.0 [18.0,29.0] | 23.0 [19.0,29.0] |
| Lung_Compliance, median [Q1,Q3] (L/ cm H2O) | 2291 | 32.0 [23.9,42.0] | 26.2 [16.0,37.0] | 28.0 [19.0,38.0] | 27.0 [18.5,36.0] |
| Minute_ventilation, median [Q1,Q3] (L/minute) | 115 | 9.5 [7.7,11.4] | 9.8 [8.2,11.5] | 9.1 [7.4,11.0] | 9.6 [7.8,11.5] |
| ABG_pH, median [Q1,Q3] | 3385 | 7.4 [7.3,7.4] | 7.4 [7.3,7.4] | 7.4 [7.3,7.5] | 7.4 [7.3,7.4] |
| ABG_PaCO2, median [Q1,Q3] (mm Hg) | 3410 | 41.0 [35.3,48.0] | 43.0 [37.0,50.8] | 42.8 [34.0,52.0] | 44.7 [37.4,52.0] |
| ABG_PaO2, median [Q1,Q3] (mm Hg) | 3377 | 87.0 [74.0,105.4] | 86.0 [72.0,102.5] | 95.9 [81.0,118.5] | 71.3 [61.9,87.7] |
| WBC_count, median [Q1,Q3] (K/UL) | 213 | 10.6 [7.3,15.9] | 13.3 [9.3,18.1] | 10.8 [7.0,16.6] | 11.2 [8.2,15.5] |
| Lymphocytes, median [Q1,Q3] (K/UL) | 2294 | 0.9 [0.5,1.4] | 0.9 [0.6,1.6] | 0.9 [0.6,1.3] | 0.8 [0.5,1.4] |
| Neutrophils, median [Q1,Q3] (K/UL) | 2215 | 8.2 [5.3,13.4] | 10.1 [6.3,14.5] | 8.7 [5.3,14.4] | 9.0 [6.2,13.1] |
| Hemoglobin, median [Q1,Q3] (g/dL) | 168 | 8.8 [7.8,10.5] | 8.5 [7.6,9.9] | 8.3 [7.5,9.6] | 9.7 [8.3,11.2] |
| Platelets, median [Q1,Q3] (K/UL) | 221 | 179.0 [91.0,276.5] | 188.0 [99.0,278.0] | 170.0 [95.0,254.0] | 202.0 [136.0,278.0] |
| Bicarbonate, median [Q1,Q3] (mmol/L) | 214 | 25.0 [22.0,28.0] | 25.0 [22.2,30.0] | 25.0 [22.0,30.0] | 25.0 [21.0,29.0] |
| Creatinine, median [Q1,Q3] (mg/dL) | 230 | 1.1 [0.7,1.7] | 1.0 [0.7,1.6] | 0.9 [0.5,1.6] | 0.9 [0.6,1.5] |
| Albumin, median [Q1,Q3] (g/dL) | 1836 | 2.7 [2.3,3.0] | 2.7 [2.4,3.0] | 2.7 [2.4,2.9] | 2.7 [2.4,3.1] |
| Bilirubin, median [Q1,Q3] (mg/dL) | 1882 | 0.7 [0.4,1.4] | 0.8 [0.4,1.9] | 0.9 [0.5,2.3] | 0.6 [0.4,0.9] |
| CRP, median [Q1,Q3] (mg/L) | 4002 | 147.0<br>[74.8,256.0] | 196.1<br>[84.7,326.7] | 158.8 [55.1,223.1] | 79.3<br>[30.7,148.9] |
| D_dimer, median [Q1,Q3] (ng/ml D-DU) | 3787 | 1732.5<br>[808.2,3368.5] | 1424.0<br>[693.5,3042.5] | 850.0<br>[453.0,2084.0] | 855.0<br>[444.0,2242.5] |
| Ferritin, median [Q1,Q3] (ng/mL) | 4727 | 734.3<br>[417.2,1306.7] | 891.8<br>[361.3,1334.4] | 426.9<br>[310.9,647.8] | 855.0<br>[582.0,1449.5] |
| LDH, median [Q1,Q3] (units/L) | 4129 | 344.5<br>[266.8,498.5] | 416.0<br>[301.0,538.0] | 307.0<br>[260.0,442.0] | 366.0<br>[297.0,491.0] |
| Lactic_acid, median [Q1,Q3] (mmol/L) | 5105 | 1.6 [1.2,2.4] | 1.7 [1.2,2.3] | 1.5 [1.1,2.3] | 1.2 [0.9,1.8] |
| Procalcitonin, median [Q1,Q3] (ng/mL) | 4851 | 0.9 [0.2,3.1] | 1.1 [0.3,4.1] | 0.9 [0.4,2.0] | 0.4 [0.1,1.3] |

**Supplemental Table 3.** Full table of all models and all imputation strategies. Extreme Gradient Boosting (XGBoost), Logistic Regression, Random Forest, LightGBM, Recurrent Neural Network (RNN), and Long Short-Term Memory (LSTM), performance on the test set, using different imputation strategies (raw, mean, and FIDDLET, detailed in Methods). Various evaluation metrics are reported to assess model performance comprehensively: AUROC (95% CI): The Area Under the Receiver Operating Characteristic Curve (AUROC), Area Under the Precision-Recall Curve (AUPRC) for intubation, Accuracy (the proportion of correctly predicted instances out of the total number of instances in the dataset and is often expressed as a percentage), F1 score (the harmonic mean of precision and recall). Class 1 indicates next-day intubated, and class 0 next-day extubated. The top models were subjected to hyperparameter tuning (bolded).

| Model Type | Imputation Method | AUROC | AUPRC | Overall Accuracy | F1 Score Class 0 | F1 Score Class 1 | Precision Class 0 | Precision Class 1 | Recall Class 0 | Recall Class 1 |
| --- | --- | --- | --- | --- | --- | --- | --- | --- | --- | --- |
| **RNN** | **binning** | **0.868**<br>(0.834, 0.900) | 0.372<br>(0.257, 0.480) | 0.714<br>(0.681, 0.745) | 0.323<br>(0.260, 0.385) | 0.818<br>(0.795, 0.842) | 0.198<br>(0.154, 0.246) | 0.985<br>(0.975, 0.995) | 0.877<br>(0.789, 0.951) | 0.700<br>(0.667, 0.733) |
| RNN | mean | 0.613<br>(0.543, 0.688) | 0.113<br>(0.079, 0.157) | 0.251<br>(0.223, 0.280) | 0.161<br>(0.126, 0.196) | 0.324<br>(0.285, 0.362) | 0.088<br>(0.068, 0.109) | 0.968<br>(0.938, 0.993) | 0.923<br>(0.852, 0.984) | 0.194<br>(0.167, 0.222) |
| **LSTM** | **binning** | **0.867**<br>(0.831, 0.899) | 0.352<br>(0.250, 0.459) | 0.709<br>(0.676, 0.741) | 0.331<br>(0.270, 0.393) | 0.814<br>(0.790, 0.837) | 0.202<br>(0.160, 0.247) | 0.991<br>(0.982, 0.998) | 0.924<br>(0.847, 0.984) | 0.691<br>(0.657, 0.724) |
| LSTM | mean | 0.666<br>(0.597, 0.728) | 0.125<br>(0.088, 0.166) | 0.541<br>(0.508, 0.578) | 0.200<br>(0.150, 0.251) | 0.678<br>(0.647, 0.709) | 0.116<br>(0.085, 0.148) | 0.959<br>(0.940, 0.976) | 0.738<br>(0.627, 0.836) | 0.525<br>(0.489, 0.560) |
| **LightGBM** | **binning** | **0.838**<br>(0.802, 0.873) | 0.242<br>(0.173, 0.314) | 0.614<br>(0.581, 0.649) | 0.278<br>(0.224, 0.330) | 0.736<br>(0.707, 0.764) | 0.163<br>(0.127, 0.199) | 0.993<br>(0.985, 1.000) | 0.953<br>(0.892, 1.000) | 0.585<br>(0.549, 0.621) |
| LightGBM | mean | 0.835<br>(0.797, 0.869) | 0.239<br>(0.173, 0.305) | 0.752<br>(0.722, 0.783) | 0.317<br>(0.247, 0.382) | 0.849<br>(0.827, 0.870) | 0.202<br>(0.152, 0.253) | 0.972<br>(0.958, 0.985) | 0.739<br>(0.630, 0.845) | 0.753<br>(0.722, 0.785) |
| **LightGBM** | **raw** | **0.839**<br>(0.802, 0.872) | 0.239<br>(0.171, 0.310) | 0.655<br>(0.621, 0.691) | 0.294<br>(0.235, 0.351) | 0.771<br>(0.744, 0.798) | 0.175<br>(0.135, 0.215) | 0.990<br>(0.980, 0.998) | 0.924<br>(0.857, 0.984) | 0.632<br>(0.596, 0.668) |
| **Logistic Regression** | **binning** | **0.853**<br>(0.808, 0.893) | 0.305<br>(0.213, 0.406) | 0.707<br>(0.676, 0.739) | 0.310<br>(0.246, 0.371) | 0.814<br>(0.790, 0.837) | 0.190<br>(0.146, 0.234) | 0.982<br>(0.970, 0.993) | 0.847<br>(0.746, 0.929) | 0.695<br>(0.663, 0.729) |
| Logistic Regression | mean | 0.797<br>(0.753, 0.839) | 0.209<br>(0.152, 0.284) | 0.652<br>(0.621, 0.685) | 0.278<br>(0.222, 0.337) | 0.771<br>(0.745, 0.796) | 0.166<br>(0.128, 0.206) | 0.982<br>(0.969, 0.992) | 0.862<br>(0.769, 0.941) | 0.635<br>(0.601, 0.668) |
| XGBoost | binning | 0.828<br>(0.786, 0.866) | 0.266<br>(0.184, 0.354) | 0.616<br>(0.581, 0.650) | 0.276<br>(0.221, 0.330) | 0.739<br>(0.709, 0.766) | 0.162<br>(0.126, 0.199) | 0.991<br>(0.981, 0.998) | 0.938<br>(0.870, 0.987) | 0.589<br>(0.552, 0.625) |
| XGBoost | mean | 0.819<br>(0.771, 0.862) | 0.230<br>(0.166, 0.302) | 0.629<br>(0.594, 0.664) | 0.259<br>(0.204, 0.314) | 0.753<br>(0.724, 0.780) | 0.153<br>(0.118, 0.192) | 0.977<br>(0.962, 0.990) | 0.831<br>(0.732, 0.921) | 0.612<br>(0.576, 0.648) |
| XGBoost | raw | 0.798<br>(0.755, 0.835) | 0.212<br>(0.151, 0.278) | 0.680<br>(0.649, 0.711) | 0.288<br>(0.228, 0.348) | 0.793<br>(0.769, 0.817) | 0.174<br>(0.133, 0.218) | 0.979<br>(0.965, 0.990) | 0.831<br>(0.733, 0.916) | 0.667<br>(0.633, 0.701) |
| Random Forest | binning | 0.835<br>(0.792, 0.873) | 0.251<br>(0.181, 0.332) | 0.666<br>(0.633, 0.700) | 0.283<br>(0.223, 0.341) | 0.782<br>(0.757, 0.809) | 0.170<br>(0.130, 0.211) | 0.980<br>(0.966, 0.992) | 0.847<br>(0.747, 0.937) | 0.650<br>(0.617, 0.686) |
| Random Forest | mean | 0.785<br>(0.725, 0.836) | 0.202<br>(0.143, 0.266) | 0.731<br>(0.702, 0.759) | 0.312<br>(0.245, 0.375) | 0.833<br>(0.810, 0.853) | 0.195<br>(0.148, 0.244) | 0.975<br>(0.961, 0.988) | 0.784<br>(0.672, 0.883) | 0.726<br>(0.695, 0.757) |

**Supplemental Table 4.**
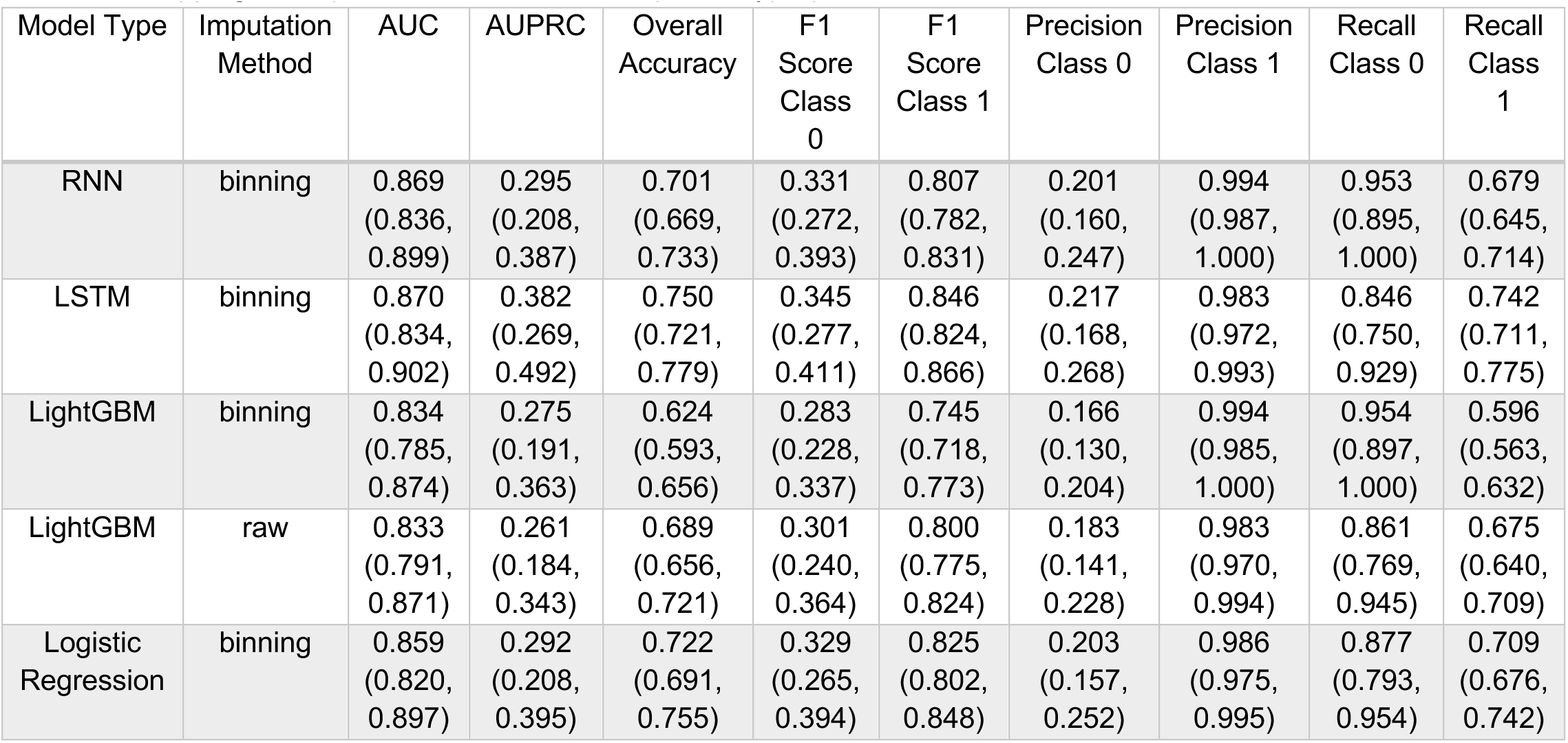
Hyperparameter optimization of top 5 performing models. Top performing models from Supplemental Table 3 (bolded) were chosen for hyperparameter optimization. The same metrics and confidence intervals from bootstrapping are reported for each model’s optimal hyperparameter set.

| Model Type | Imputation Method | AUC | AUPRC | Overall Accuracy | F1 Score Class 0 | F1 Score Class 1 | Precision Class 0 | Precision Class 1 | Recall Class 0 | Recall Class 1 |
| --- | --- | --- | --- | --- | --- | --- | --- | --- | --- | --- |
| RNN | binning | 0.869<br>(0.836, 0.899) | 0.295<br>(0.208, 0.387) | 0.701<br>(0.669, 0.733) | 0.331<br>(0.272, 0.393) | 0.807<br>(0.782, 0.831) | 0.201<br>(0.160, 0.247) | 0.994<br>(0.987, 1.000) | 0.953<br>(0.895, 1.000) | 0.679<br>(0.645, 0.714) |
| LSTM | binning | 0.870<br>(0.834, 0.902) | 0.382<br>(0.269, 0.492) | 0.750<br>(0.721, 0.779) | 0.345<br>(0.277, 0.411) | 0.846<br>(0.824, 0.866) | 0.217<br>(0.168, 0.268) | 0.983<br>(0.972, 0.993) | 0.846<br>(0.750, 0.929) | 0.742<br>(0.711, 0.775) |
| LightGBM | binning | 0.834<br>(0.785, 0.874) | 0.275<br>(0.191, 0.363) | 0.624<br>(0.593, 0.656) | 0.283<br>(0.228, 0.337) | 0.745<br>(0.718, 0.773) | 0.166<br>(0.130, 0.204) | 0.994<br>(0.985, 1.000) | 0.954<br>(0.897, 1.000) | 0.596<br>(0.563, 0.632) |
| LightGBM | raw | 0.833<br>(0.791, 0.871) | 0.261<br>(0.184, 0.343) | 0.689<br>(0.656, 0.721) | 0.301<br>(0.240, 0.364) | 0.800<br>(0.775, 0.824) | 0.183<br>(0.141, 0.228) | 0.983<br>(0.970, 0.994) | 0.861<br>(0.769, 0.945) | 0.675<br>(0.640, 0.709) |
| Logistic Regression | binning | 0.859<br>(0.820, 0.897) | 0.292<br>(0.208, 0.395) | 0.722<br>(0.691, 0.755) | 0.329<br>(0.265, 0.394) | 0.825<br>(0.802, 0.848) | 0.203<br>(0.157, 0.252) | 0.986<br>(0.975, 0.995) | 0.877<br>(0.793, 0.954) | 0.709<br>(0.676, 0.742) |

**Supplemental Table 5:** Hyperparameter search space for fine-tuning RNN, LSTM, LightGBM and logistic regression models. Top hyperparameter combinations for each model was chosen by AUROC.

| Model | Imputation | Hyperparameters | # of Models |
| --- | --- | --- | --- |
| LSTM | Binning | learning_rate: {0.001, 0.01, 0.1} * unit_1: {32, 64, 128} * unit_2: {16, 32, 64} * batch_size: {32, 64, 128} | 81 |
| RNN | Binning | learning_rate: {0.001, 0.01, 0.1} * unit_1: {32, 64, 128} * unit_2: {16, 32, 64} * batch_size: {32, 64, 128} | 81 |
| Logistic Regression | Binning | C:{0.01, 0.1, 1} * penalty: {l1, l2} * solver: {liblinear} | 6 |
| LightGBM | Binning | num_leaves:{31, 61, 127} * learning_rate:{0.01, .05, .1} * n_estimators: {100, 200, 500} | 27 |
| LightGBM | Raw | num_leaves:{31, 61, 127} * learning_rate:{0.01, .05, .1} * n_estimators: {100, 200, 500} | 27 |
| XGBoost | Raw | n_iterations: {1000} * max_depth: {30} | 1 |
| XGBoost | Binning | n_iterations: {1000} * max_depth: {30} | 1 |
| Random Forest | Binning | default | 1 |

## Supplemental Documents

### NU SCRIPT Study Investigators

TRIPOD Checklist [Moons KG, Altman DG, Reitsma JB, Ioannidis JP, Macaskill P, Steyerberg EW, Vickers AJ, Ransohoff DF, Collins GS. Transparent Reporting of a multivariable prediction model for Individual Prognosis Or Diagnosis (TRIPOD): Explanation and Elaboration. Ann Intern Med. 2015;162(1):W1-W73. PMID: 25560730]

